# Adults’ experiences of comorbid insomnia and pain: “Entangled vicious cycle” driven by four primary factors

**DOI:** 10.64898/2026.09.29.26364320

**Authors:** Adalbjorg Albertsdottir, Anna Sigridur Islind, Hafdis Skuladottir, Sigridur Halldorsdottir, Kristin Anna Olafsdottir, Erla Bjornsdottir, Arnor Vikingsson, Gabriel Natan Pires, Erna Sif Arnardottir

## Abstract

Chronic pain and insomnia are common, each has serious consequences and they often coexist [1–5]. This novel study employed a phenomenological method to explore adults lived experiences of the core and meaning of comorbid insomnia and pain, here conceptualized as COMIP. The empirical data is based on 22 semi-structured interviews with adults with COMIP. Each interview was analysed, developing themes and analytical frameworks for all participants, which were then compared to construct one main analytical framework. The consolidated criteria for reporting qualitative research were used [6].

The essence of COMIP was: A vicious cycle of insomnia and pain driven by four intertwined primary factors with cross effects, which reduced sleep quality and quality of life. Four main themes emerged: Firstly, insomnia with hyperarousal like racing thoughts, and difficulties with finding comfortable sleeping positions due to pain, disturbed sleep and decreased sleep quality. Secondly, daytime symptoms like reduced energy, with reduced daytime function and reduced quality of life. Thirdly, 24-hour hyperarousal, stress and distressing experiences such as suffering. Finally, contextual factors like treatment reduced and/or increased COMIP.

The main contribution of this research is three-fold; mapping of the experiences of living with COMIP, the conceptualization of COMIP, and introduction of novel concepts. Our findings indicate that, when evaluating patients with COMIP, there is value in exploring the four COMIP factors. Although, it varies between individuals which factor/s is central to their COMIP experience, we emphasize that COMIP is distressing, and patients are likely to need emotional support and validation.

## 1. Introduction

The main diagnostic criteria for chronic insomnia include difficulties initiating or maintaining sleep, accompanied by daytime symptoms such as fatigue. These symptoms must occur at least three times per week for a minimum of three months and should not be better explained by another condition [7]. From this point on, chronic insomnia will be referred to as insomnia. The prevalence of insomnia in the general population is high, recent meta-analyses estimate 25,1% of the population experience insomnia symptoms and the prevalence of chronic insomnia to range from 12.4% [1] to 16.2% [8].

The diagnostic criteria for chronic pain includes the presence of pain for at least three months, associated with significant emotional distress and/or functional disability and not better accounted for by another condition [9]. Examples of chronic pain include pelvic pain, fibromyalgia and musculoskeletal pain [9]. From this point on, chronic pain will be referred to as pain. The prevalence of pain in the general population is estimated to range from 20.3% [10] to 24.3% [2].

Insomnia and pain have both been linked to adverse health outcomes [3,4,11], reduced quality of life [4,12] and economic burden for individuals and society [12,13]. For example, chronic musculoskeletal pain has been identified as one of the most common reasons for disability [14].

A significant comorbidity between insomnia and pain has been established. A cross-sectional community-based study found that 50% of participants with insomnia had pain, compared to only 18% of those without insomnia [5]. Conversely 49% of participants with pain had insomnia, while only 17% of participants without pain had insomnia [5]. A systematic review with meta-analysis found even stronger comorbidity between insomnia and pain, with 73% of patients with pain experiencing insomnia [15]. Santos and colleagues examined the bidirectional relationship between sleep problems and chronic musculoskeletal pain in a systematic review with meta-analysis [16]. Their study provided evidence of a longitudinal association between sleep problems and incidence-persistence of chronic musculoskeletal pain in adults as well as a bidirectional relationship between the two. However, the nature of this bidirectional relationship requires further investigation, and we concur with Arnardottir and colleagues [17] that in sleep science the experience of people is equally important as quantitative measurements. Considering the prevalence, effects and comorbidity of insomnia and pain, the importance of a deeper understanding of the lived experiences of comorbid insomnia and pain cannot be overstated.

Naming a phenomenon facilitates shared understanding and further exploration of it. Sweetman and colleagues coined the term COMISA (comorbid insomnia and sleep apnoea) [18]. Although it has long been recognized that insomnia and sleep apnoea are often comorbid, we now have a shared language for this specific phenomenon. By following in the footsteps of Sweetman and colleagues’ [18,19] we propose the term COMIP for comorbid insomnia and pain, a term which cannot be seen as favouring one over the other, see Figure 1.

**Figure 1.**
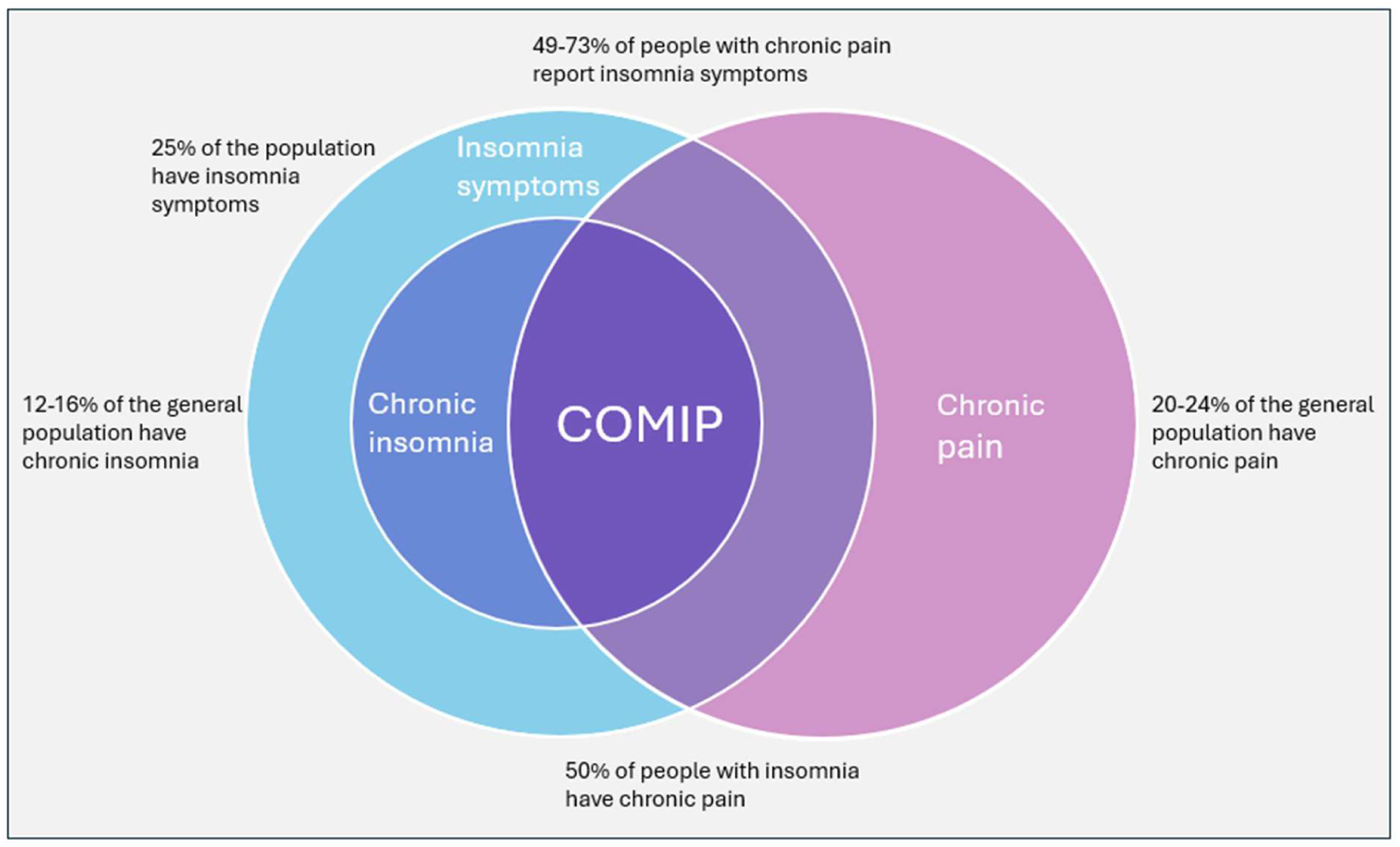
Significant comorbidity between insomnia and pain has been established, their overlap is defined as COMIP. Taylor and colleagues found that pain was significantly more common in those with insomnia than without (50% vs. 18%), and insomnia was much higher in those with pain than without (49% vs. 17%) [5]. Numbers in the figure are from several studies [1,2,5,8,10,15].

Despite the high prevalence of insomnia and pain, and the significant comorbidity between them, there is a lack of qualitative studies exploring COMIP as a phenomenon in itself. Although, a qualitative study from 2014 came close; the aim was to explore insomnia in participants with chronic, work-related musculoskeletal pain. Participants described sleep problems consistent with insomnia, which exerted a significant impact on their lives. They attributed their insomnia to psychological stress like racing thoughts and the physical stress of pain causing e.g. difficulties in finding a comfortable sleeping position [20]. In line with this, a phenomenological study of people in rehabilitation with chronic musculoskeletal pain found similar results regarding the participants’ sleep. Participants experienced difficulties falling and staying asleep because of pain and hyperarousal symptoms like worries and anxiety [21].

However, like most qualitative papers that cover insomnia and pain, one was the main topic and the other one a sidenote. Taking this even further, prominent qualitative reviews covering insomnia on one hand and pain on the other, do not mention the other [22–24] despite the hight comorbidity and bidirectional impact [5,16].

Pain research generally resumes a biopsychosocial framework, acknowledging that pain is not “somatic” or “psychogenic” but has multiple interacting contributors [9]. The theoretical framework of this paper is the International Classification of Functioning, Disability and Health (ICF) [25]. Thereby, we ensure meticulous attention not only to biopsychosocial factors, but also to personal and environmental factors, in the context of function, disability and health.

Given the gap in the literature regarding lived experiences of those with COMIP, a phenomenological approach was chosen with the aim to explore the essence of the COMIP experience. The research question was twofold: What are the participants lived experiences of the core, and the meaning, of COMIP? Before commencing the study, our preunderstanding was that COMIP would be characterized by a vicious cycle of insomnia, pain, and hyperarousal. The Vancouver-School of Doing Phenomenology (hereinafter referred to as the Vancouver-School) was selected to answer the twofold research question. In the Vancouver-School each participant is seen as a case-study, with deep analysis of individual cases. Then followed by inter-case analysis and systematic reconstruction for the overall results [26].

The main contribution of this paper is threefold. Firstly, we provide rich descriptions of the participants lived experiences of COMIP. Secondly, we introduce novel concepts which emerged through phenomenological analysis of the empirical data and finally, we conceptualise COMIP as a serious condition requiring attention in research and healthcare.

## 2. Methods

The aim of the study was a deeper understanding of adults lived experiences of COMIP, with the purpose to facilitate increased awareness, understanding and knowledge about COMIP to advance research and health care services for people with COMIP. To reach this aim, phenomenological methodology with purposeful sampling was employed.

### 2.1. Participants and selection criteria

The research population was Icelandic speaking adults aged 18 to 67 years, of all genders from all over the country, with more than three months experience of COMIP that disturbed sleep three times or more per week. Before commencing the study, it was approved by the National Bioethics Committee and Data Protection Agency of Iceland, approval number VSN2501010. A purposive sample of eleven participants was recruited through advertising on social media, and in collaboration with Iceland largest rehabilitation hospital, Reykjalundur. The participants were between 25 and 65 years old, and the majority were women who saw the study advertised online. The participants signed an informed consent and answered screening questionnaires with selection criteria through a personalized link in REDCap (Research Electronic Data Capture) a secure, web-based software platform, supporting data capture in research studies [27]. The inclusion criteria were: Pittsburgh Sleep Quality Index (PSQI) score > 5 (scale from 0-21, score >5= indicates poor sleep quality) [28], Insomnia Severity Index (ISI) score ≥ 10 (scale from 0-28, score ≥10 indicates insomnia) [29,30] and pain intensity was measured on the numerical rating scale (NRS) (scale from 0-10, 0=no pain and 10=the worst pain possible) [9]. No minimal pain value was used for study inclusion, but pain needed to disturbed sleep ≥ 3 times per week for more than three months. The exclusion criteria were STOP-Bang ≥ 3 (scale from 0-8, score ≥3 indicates intermediate risk for sleep apnoea) [31], fulfilling the diagnostic criteria of restless legs syndrome [32] or self-reported diagnosis of sleep apnoea, restless legs syndrome or serious comorbid health condition, such as cancer or neurodegenerative diseases. The Epworth Sleepiness Scale (ESS) (scale from 1-24, score >10 indicates excessive daytime sleepiness) was used to enhance understanding of the participants’ sleepiness but was not used for the inclusion-exclusion criteria [33].

All participants scored 12 or higher on the ISI and 8 or higher on the PSQI, which aligns with the selection criteria and supports a diagnosis of clinical insomnia and poor sleep quality. Average pain intensity on the NRS over the past three months was 6, see Table 1. To protect the identity of participants, they chose a pseudonym and age is described in five-year intervals.

**Table 1.** Overview of the 11 participants, their characteristics and screening questionnaire results.

| Pseudonym | Age range (yrs) | Gender | ISI | NRS* | PSQI | ESS | STOP BANG | Type of chronic pain |
| --- | --- | --- | --- | --- | --- | --- | --- | --- |
| Adam | 40-45 | M | 22 | 8 | 13 | 6 | 2 | Musculoskeletal pain |
| Anna | 35-40 | F | 17 | 6 | 16 | 15 | 1 | Fibromyalgia and musculoskeletal pain |
| Aria | 55-60 | F | 12 | 5+ | 16 | 11 | 1 | Fibromyalgia |
| Ella | 30-35 | F | 15 | 6-7 | 11 | 6 | 0 | Fibromyalgia |
| Iris | 45-50 | F | 22 | 7 | 13 | 19 | 2 | Fibromyalgia |
| Ivy | 35-40 | F | 23 | 4-6 | 18 | 6 | 2 | Fibromyalgia, musculoskeletal pain and endometriosis |
| Magnus | 45-50 | M | 25 | 5 | 15 | 0 | 1 | Musculoskeletal pain |
| Sage | 40-45 | F | 27 | 6-9 | 19 | 18 | 1 | Fibromyalgia and musculoskeletal pain |
| Sarah | 25-30 | F | 23 | 7 | 16 | 17 | 2 | Fibromyalgia and musculoskeletal pain |
| Sophie | 45-50 | F | 20 | 7 | 16 | 17 | 2 | Fibromyalgia |
| Tristan | 60-65 | M | 13 | 5 | 8 | 11 | 1 | Musculoskeletal pain |
| MEAN ± SD | 43 | | 19.9±4.7 | 6.0±1.1 | 14.7±3.0 | 11.4±6.0 | 1.3±0.7 | |
| RANGE | 25-65 | M: 3<br>F: 8 | 12-27 | 4-9 | 8-19 | 0-19 | 0-2 |  |

Abbreviations: M (male), F (female), ISI (Insomnia Severity Index), NRS (Numerical rating scale), PSQI (Pittsburgh Sleep Quality Index), ESS (Epworth Sleepiness Scale), STOP-Bang (STOP-Bang questionnaire), SD (standard deviation).
\* The question measuring pain intensity was: What was your average pain intensity over the past three months on a scale of 0-10, with 0 being no pain and 10 being the worst pain possible? Some participants answered that question using a range, rather than with a single value.

### 2.2. The methodology

As a research method, phenomenology can be used to describe the shared essence of a concept and define concepts, building on descriptions from all the participants lived experiences [34]. The methodology used in this study is the Vancouver-School, the epistemological stance of which is interpretivist/constructivist phenomenology, primarily influenced by Spiegelberg (phenomenology), Ricoeur (hermeneutics/interpretivist) and Schwandt (constructivist/interpretivist)[26,35–38]. A key strength of this methodology is its structured approach to case construction. By deconstructing individual interviews into key themes and then reconstructing all the interviews into a cohesive whole, the process ensures both depth and synthesis of newfound knowledge. The four-fold verification which is part of the research process is also a strength [26]. An overview of the systematic research process in this study is described in Table 2.

**Table 2.** The research process of the Vancouver-School in this study. Adapted from Halldorsdottir [26].

| **Steps of the research process** | **What was done in the present study** |
| --- | --- |
| *Step 1. Selecting dialogue partners (the sample)* | Fifteen participants signed the informed consent and answered the selection criteria questionnaires. Eleven participants met the selection criteria, eight women and three men. |
| *Step 2. Silence (before entering a dialogue)* | The primary researcher reflected on her preconceived ideas, wrote them down and consciously set them aside as much as possible. The researcher is a registered nurse with 15 years of rehabilitation experience, MSc in Health Science and a PhD student in Sleep Science. |
| *Step 3. Participating in a dialogue (data collection) Verification 1* | Two interviews were conducted with each participant, a total of 22 interviews. The average interview length was 40 minutes. The researcher conducted all the interviews, which were recorded on Teams, written verbatim in a computer, and encrypted. First verification was within each dialogue situation, where the researcher verified her understanding with the participant. |
| *Step 4. Sharpened awareness of words (beginning data analysis)* | Data analysis started in the interviews, so data collection and data analysis ran concurrently. The interviews were then read repeatedly and iteratively to identify the essence of each interview, trying to answer the research questions for that participant. |
| *Step 5. Beginning consideration of essences (coding)* | Each interview was analysed further, and data was organized into main themes and subthemes to begin constructing the essence of the lived experience of COMIP for that participant. |
| *Step 6. Constructing the essential structure of COMIP from each case (individual case construction)* | The main themes and subthemes in each participant's story were analysed, and the most significant themes were built into an individual analytical framework. |
| *Step 7. Verifying each case construction with the relevant participant. Verification 2* | In the second interview each participant verified his or her analytical framework, constructed by the first researcher and elaborated on the findings. |
| *Step 8. Constructing the essential structure of COMIP from all the cases (metasynthesis of all the different case constructions)* | All individual analytical frameworks were compared to construct one main analytical framework. In this final data analysis, five more researchers stepped in, reflected on the data, and together reconstructed part of the preliminary findings. Such triangulation is encouraged in the Vancouver-School. |
| *Step 9. Comparing the essential structure of COMIP with the research data for verification. Verification 3* | For verification, the researcher re-read all the interviews and compared them to the final analytical framework. |
| *Step 10. Identifying the overriding theme describing COMIP (construction of the main theme)* | COMIP was a vicious cycle driven by four intertwined primary factors: I. The pain/insomnia sleep disturbance II. Daytime symptoms III. Hyperarousal, stress and distress IV. Contextual factors, like treatment and environment. |
| *Step 11. Verifying the essential structure with the research participants Verification 4* | The overall findings were presented by the researcher to two participants. This is part of the verification process. They both agreed with and verified the findings. |
| *Step 12. Writing the findings (multivoiced reconstruction)* | The participants are quoted directly to increase the credibility and trustworthiness of the findings and conclusions. The first draft was written by the researcher, but all authors contributed to rewriting it. |

During the analysis process, the researchers go through seven main cognitive stages in a circular process that is repeated across all the research steps (Figure 2). This is not a linear process but a repetitive, iterative one, in which the interpretive circle is used in all the steps [26].

**Figure 2.**
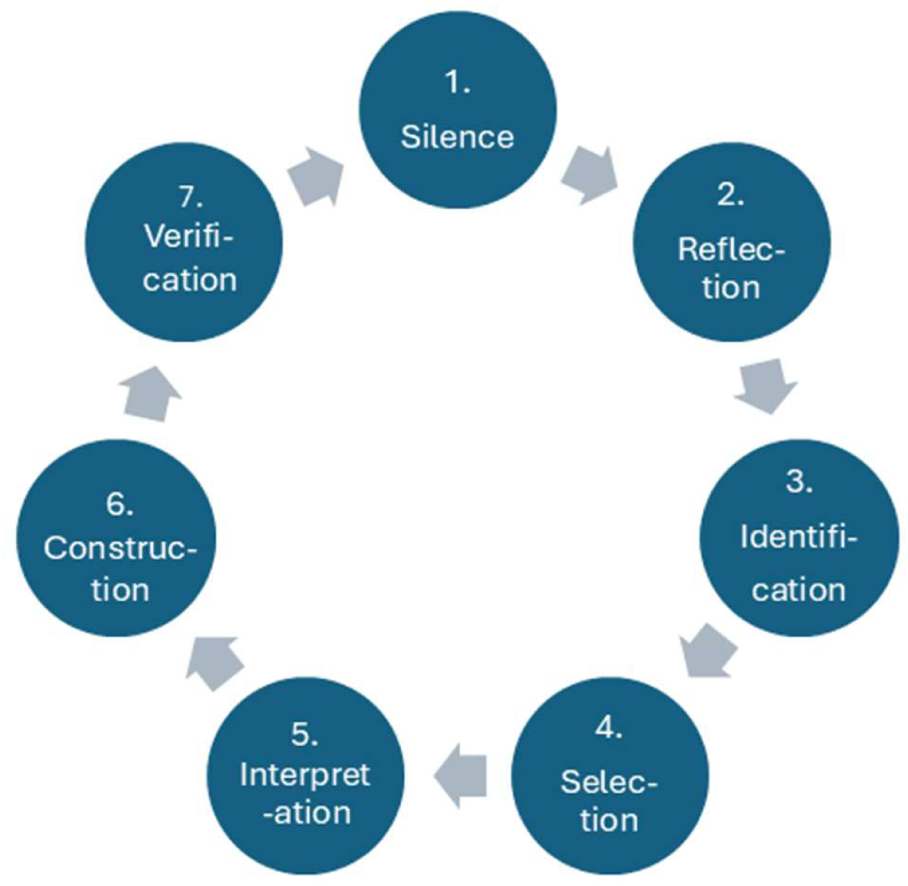
The repetitive cycle of the seven stages in the Vancouver-School. Repeated in all 12 steps of the Vancouver-School. Adapted from Halldorsdottir[26].

In the Vancouver-School, all participants’ voices are to be heard, and data saturation can be reached with five to fifteen participants, preferably each interviewed twice, and a minimum of 10 interviews [26]. In this study, data saturation was reached after 20 interviews, but two more interviews were conducted to confirm it. Therefore, all participants were interviewed twice. The interview frame is provided in Appendix A. The interviews were recorded, transcribed verbatim and analysed using the Vancouver-School method outlined above. To increase trustworthiness, six researchers participated in research triangulation (Step 8 in Table 2), and the consolidated criteria for reporting qualitative research were followed [6].

## 3. Results

### 3.1. The essence of the experience

The overarching theme: COMIP was a vicious cycle of insomnia and pain driven by four intertwined primary factors with cross effects, which reduced sleep quality and quality of life. It varied between individuals which primary factor was central to their experience. The four main themes which emerged are the four primary factors, identified as the building blocks of the COMIP experience. The main themes and their intertwined nature are shown in Figure 3.

**Figure 3.**
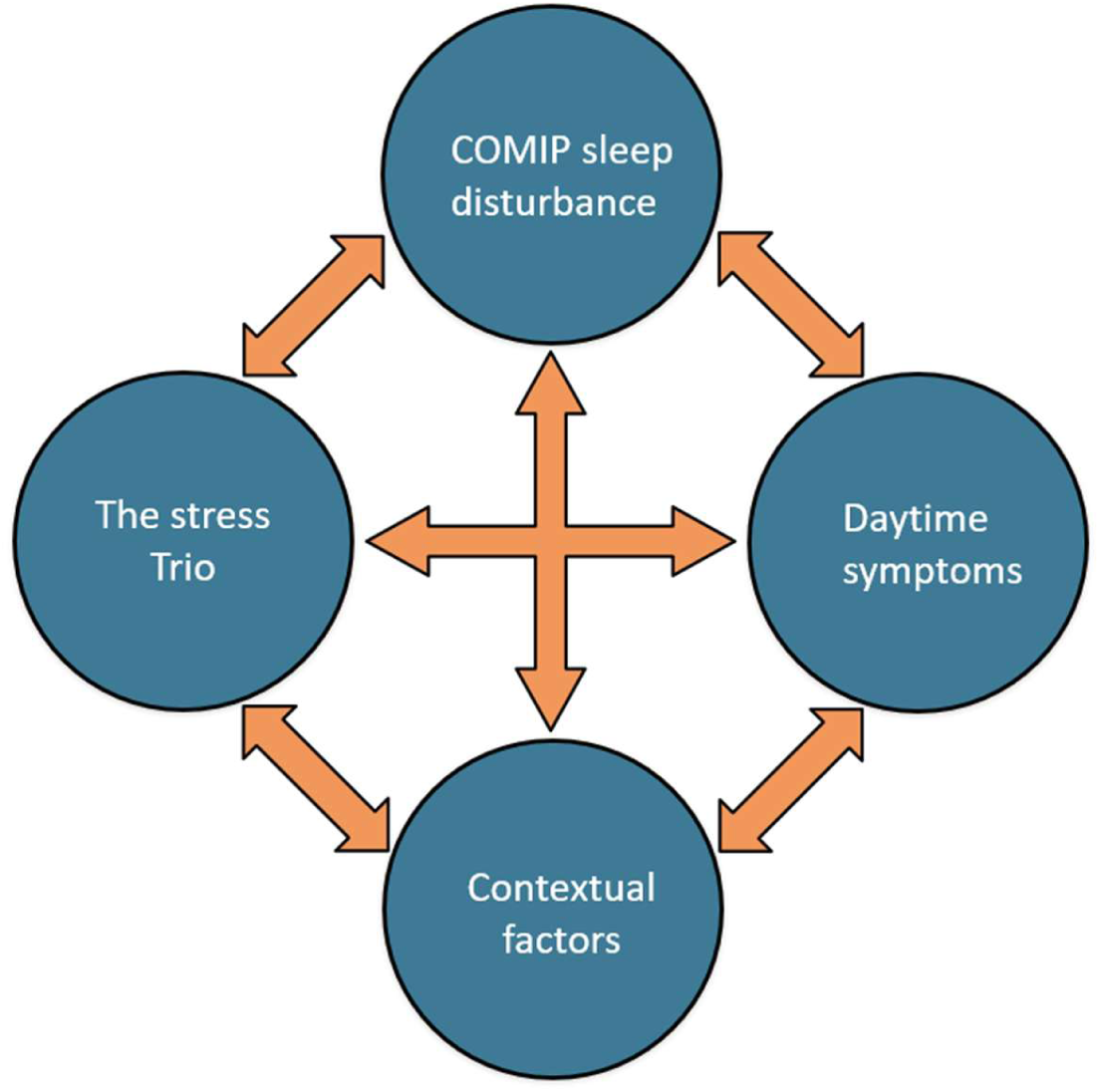
The COMIP experience: A vicious cycle of insomnia and pain driven by four intertwined primary factors.

I. In the COMIP sleep disturbance, the pain and insomnia magnified each other and could not be clearly separated. Amongst the effects of the COMIP sleep disturbance was reduced ability to stay asleep and impaired sleep quality. II. Increased daytime symptoms were part of the cycle, particularly fatigue, which led to reduced ability to participate in work and social life, and a decline in quality of life. III. In the stress trio; 24-hour hyperarousal, stress and distressing emotions such as hopelessness, were reciprocally related to the other COMIP factors. IV. Finally, contextual factors like treatment and environment were found to both alleviate and aggravate the COMIP experience. It varied between individuals which primary factor was central to their experience.

3.2. The COMIP sleep disturbance with decreased sleep function and sleep quality: *“An attack-like interplay of pain and insomnia…*”

Participants experienced insomnia with hyperarousal like racing thoughts, alongside difficulties in finding comfortable sleeping positions and awakenings due to pain. The insomnia and pain magnified one another; they could not be clearly separated and caused reduced ability to sleep with reduced sleep quality. The experience of this combined sleep disturbance was severely distressing; it was described as “a marriage from hell” (Adam) and the nature of this relationship was captured as: *“An attack-like interplay of pain and insomnia…*” (Sage). We refer to the combined experience of insomnia and pain disturbing sleep as the COMIP sleep disturbance.

3.2.1. The experience of insomnia in COMIP was characterized by hyperarousal symptoms that were increased by pain: *“it’s distressing to know that you’re going to wake up at night because of pain”* (Iris), and stress: *“if there is a lot of stress during the day, I sleep worse at night.”* (Ella).

3.2.2. The experience of pain as a sleep disturbance in COMIP was unanimously described as difficulties with finding comfortable sleeping positions and/or awakenings due to pain: *“…hard for me to find a comfortable sleeping position…difficulties turning in bed and then wake up because of the pain.”* (Tristan).

3.2.3. In the COMIP sleep disturbance the pain sleep disturbance (difficulties finding comfortable sleeping positions and awakenings, due to pain) and insomnia (characterized by hyperarousal symptoms) magnified one another and could not be clearly separated, it was described as an: “…entangled vicious cycle…” (Sophie) and a: “…*really bad combo, waking up because of the pain and having such a hard time falling asleep because of your head* [insomnia symptom]. *This is such a cycle, such a circular nonsense, with cross-effects, back and forth.”* (Adam).

3.2.4. Experiences of reduced sleep function with reduced sleep quality: Participants described feeling like the ability to sleep was under attack by their own body: *“An attack-like interplay of pain and insomnia…*” (Sage). It was described how the COMIP sleep disturbance adversely affected sleep by impairing the ability to sleep and by disrupting sleep architecture. Specifically, participants described that COMIP caused difficulties falling and staying asleep, short sleep duration, and shallow, non-restorative sleep. With reference to the participants descriptions: “…*your own body somehow attacking this function*” (Sarah), we refer to these experiences as reduced sleep function, composed of five subthemes, which are presented with verbatim quates in Table 3. The participants experiences of poor sleep were supported by the mean PSQI score of 14,7 (Table 1).

**Table 3.** Participants’ descriptions of decreased sleep function, characterized by decreased ability to sleep alongside reduced sleep depth and non-restorative sleep.

| Subthemes of decreased sleep function | Description of theme | Quotes from participants |
| --- | --- | --- |
| Difficulties falling asleep | Participants described how both the physical discomfort of the pain and the hyperarousal symptoms of the insomnia reduced their ability to fall asleep | *"I often have difficulty falling asleep, my head is all over the place accompanied with tension and pain...it's also hard to get into a comfortable sleeping position."* (Sage) |
| Difficulties staying asleep | Difficulties staying asleep because of COMIP were consistently reported by participants | *"...always waking up and turning, changing positions...wake up feeling very uncomfortable"* (Ella)<br>*"It's kind of like you ... don't have the capacity to stay asleep. It's like a lack of ability to fall asleep and stay asleep."* (Ivy) |
| Short sleep | Short sleep duration (range 4.0-6.5 hours) was experienced by nine* participants and contributed to the distressing experience of COMIP | [Magnus's sleep problems were characterized by short sleep which was really distressing] *"...this is just one of the hardest things I've ever had to deal with. I just find it really hard... I'm not getting enough sleep, and it's been like that for so long."* (Magnus). |
| Shallow and unrestful sleep | Participants described shallow, unrestful sleep and being easily awakened. | *"...the sleep is shallow and unrestful...waking up at least every hour and then I'm trying to find a position to feel less pain here, or there...it's such a mix, I can't tell the difference. [between the pain and the insomnia]"* (Aria)<br>*"...the sleep is also shallow, everything wakes me...When I then wake up, I feel like I haven't slept at all"* (Sage) |
| Non-restorative sleep | Participants consistently described non-restorative sleep. Specifically, most of them reported waking up just as tired as they had been when they went to sleep. | *"Sleep without rest...you go to bed tired and you wake up just as tired."* (Iris) |

\* Two of the participants reached sleep duration of 7.0-7.5 hours

3.3. Daytime symptoms with decreased; daytime function and quality of life: *“My life has shrunk a lot”*

Participants experienced daytime symptoms such as pain, fatigue and brain fog; However, they emphasized the impact of their lack of energy, which affected their daytime function and quality of life.

3.3.1. All participants described having daytime symptoms such as fatigue and reduced energy, increased pain and stiffness, worse mental health, and cognitive symptoms like reduced concentration and brain fog. Specifically, Tristan described: *“…profound lack of energy. You’ve only got a small window of energy before it runs out.”* (Tristan), Adam felt the same and described it as: *“…feeling like a ghost…”* (Adam) and Sophie described how brain fog affected daily life: *„…the brain fog gets much worse, you completely forget what spoon is. You’re asking the kids to fetch: Oh, the thing you use to eat yogurt with.*” (Sophie)

3.3.2. The daytime symptoms, especially the lack of energy affected daytime function with effects on relationships, social life, work and quality of life: *„I think this mostly affects what I have energy to do during the day, I think it has the biggest impact there, because it affects social life and work.“* (Sarah). *“…I had a job and I had an active social life, my life has shrunk a lot.”* (Anna).

I’m much more limited than I was, I can’t go camping with the kids anymore. I can do a lot less than I used to…You’re always trying to find the energy to do what you want to do, but then you usually pay for it, just by being exhausted for the next few days. It’s just something you have to weigh up. It affects everything and limits your quality of life. (Sophie).

4. The stress trio: 24-hour hyperarousal, stress and distress - *“Everything is simmering”*

Three closely interwoven states; hyperarousal, stress, and distress, were described in varying combinations by participants. They ran like a red thread through the COMIP experience. Rather than presenting as separate or sequential experiences, they were described as continuously active and mutually reinforcing; each intensified the others, and together they amplified the other COMIP factors, which in turn fed back into further hyperarousal, stress, and distress.

Participants emphasized this reciprocal, self-perpetuating quality, captured vividly in one participant’s description of the experience as “everything is simmering.” We refer to these intertwined experiences as the stress trio.

4.1. Descriptions of hyperarousal like “fight or flight” and an “*anxious-stressful feeling*”: *“…I often wake up, and then I get this restless feeling, I just, no, okay, I can’t fall asleep, and then I get this anxious-stressful feeling.”* (Sage). Hyperarousal was also described as fight or flight, and it was connected to sleep problems and stress:

> *Constantly in fight or flight mode and then it goes into a vicious cycle…you can’t calm the system, you can’t recover, neither in mind, sleep nor body because everything is simmering everywhere…When I slept badly, I would have those thoughts, I have to sleep to be able to wake up, I can’t be awake, I have to, I have to. And having to be able to sleep to be able to deal with this [had been bullied at work] the next day, and then of course stress, what can I expect tomorrow? (Aria)*

4.2. The 24-hour effects of stress in COMIP were vividly described: *“I feel stressed from lack of rest, which then causes stress at work and at home.” (Magnus)*.

> *Describe COMIP as one concept, just stress!!…when the pain is interfering with sleep, and you sleep worse, you are more sore and tired during the day, which often causes a lot of stress, and I find that if there is a lot of stress during the day, I sleep worse at night. (Ella)*

4.3. Distress was evident by descriptions of difficult emotions during night and day, such as giving up, hopelessness, exhaustion, worries, misery and suffering. Ivy wiped tears when she was describing the distressing experience of not being able to sleep: *“…it’s suffering, it’s that and hopelessness. I get so exhausted sometimes, you see?“* (Ivy).

5. Contextual factors: “I think this might be related to…”

At the end of each interview, the participants were asked if there was anything else they wanted to add. This led to the identification of contextual factors, a primary factor that bidirectionally influenced participants’ sleep. This main theme had five sub-themes: treatment, environment, other sleep problems, personal and social factors. It was the only main theme that could not only increase COMIP but also decrease it. The sub-themes are described in Table 4.

**Table 4.** Participants described contextual factors, which reduced and/or increased COMIP.

| Subthemes of contextual factors | Description of the subtheme | Examples from participants |
| --- | --- | --- |
| Treatment burden vs. results. | All participants had sought treatment for their sleep problem. Treatment often had minimal effects, and the same intervention could be helpful or unhelpful and very burdensome. | An example of this is a course from the same provider, digital cognitive behavioural therapy for insomnia.<br>For Ella this course was helpful: “[n.n.] was a really great course.” “You learn a lot there” Ella managed to stop taking sleeping pills and adopt methods from the course.<br>For Magnus the same course was a very distressing experience, specifically the sleep restriction part: “...and when you have been doing this for a few days in a row. It is just really extreme [strong emphasis on extreme] and this is just the most difficult thing I have ever experienced...this just didn’t work...” |
| Environment magnified and attenuated COMIP. | The importance of a comfortable and safe environment to achieve restful sleep was identified.<br>Environmental factors that disturbed sleep were also identified, like ambient noise and low weather pressure. | The importance of a comfortable bedroom environment: “...it’s really important to create a bedroom that’s just for sleeping...to create an oasis for myself, to have the right temperature.” (Anna)<br>The importance of a safe environment: “...I never knew what to expect. Always in fight or flight mode and then it goes into a vicious cycle. I feel like you can say, a safe environment has such a huge impact on sleep and pain, then you achieve a completely different kind of rest.” (Aria)<br>Ambient noise could be very disturbing: “...the underfloor heating in the garage... Even if it was really low, then... [I] can’t fall asleep with it...” (Sage)<br>For Ivy the weather influenced both pain and sleep: “...I’m just like a weather station, when there’s low pressure weather over the country, it really impacts me. |
| Personal factors could simultaneously be helpful and hindering. | Personal factors like being solution oriented and resilient, influenced participants behaviour. | All participants had sought solutions/help for their sleep problems and resilience was manifested in relation to daytime function. Only three of the participants were still working and it came with severe challenges like falling |
|  | While being solution oriented some participants simultaneously experienced self doubt or lack of insight.<br>Participants described the need to control one 's own perspective, to continue with their daily life despite COMIP. | asleep under the wheel. Although it was also hard to function in daily life despite not having a job, the participants showed resilience: *"...I had a job and I had an active social life, my life has shrunk a lot. But I'm increasing it again and I'm not giving up."* (Anna).<br>Experiencing self-doubt or lack of insight: *"The thing is, you're just so used to your own condition, that you're terrible at judging it"* (Sarah).<br>Controlling your perspective: *"...when you think about it Pollyanna was being abusive towards herself to get through life...And that's really just what you have to do, or you know, with gentleness of course."* (Ivy). |
| Social factors bidirectionally influenced COMIP. | COMIP reduced the ability to participate in work and social life, while work and social life sometimes increased COMIP.<br>Interpersonal validation of the COMIP experience was accompanied with relief. It was expressed that COMIP being validated as a shared experience was accompanied with a good feeling. | Of the 11 participants, only three were still working: *"There are a lot of things that I can't participate in anymore, because of fatigue and lack of energy, because I don't sleep well enough. Like just going to social events, or you know, meeting people. I'm no longer working, haven't been for almost two years because of all this sleep issues..."* (Ivy)<br>Social life, work and personal relationships also influenced COMIP. Aria had been bullied at work, which magnified stress and increased her COMIP. Regarding close personal relationships, Sage described helpful support and understanding from her husband during sleepless nights, while another participant described that her husband didn't seem to understand how vulnerable she was to interruptions like snoring, at night.<br>*"Sometimes I feel like I am a bit strange just that sleep is such a big deal..." [later in the interview Aria described feeling better, knowing that this was a shared experience] "...so good to know [emphasis on good to know] that you are not alone and that this is kind of synonymous. You are not crazy, that there are just some other people saying the same things"* (Aria). |
| Other sleep problems also affected sleep. | In addition to COMIP, other factors were identified as sleep problems that affected participants' sleep. Such as nocturia and nightmares, and factors like attention deficit hyperactivity disorder (ADHD) and the menstrual cycle. | Other sleep problems that were described were nocturia and nightmares, and four participants described a possible link between ADHD or being on the spectrum, and their sleep problem:<br>*"...I've had trouble sleeping since I was a little child... I was a bedwetter and when I didn't wet the bed I would still often wake up and have insomnia at night. But I think this [sleep problem] might be related to ADHD, autism spectrum because it also runs in my family..."* (Ivy).<br>The menstrual cycle was also identified spontaneously as influencing sleep:<br>*"I've been experiencing it so much lately that the menstrual cycle has a lot to do with it too. Just wanted to throw that in...That is, the period between ovulation and menstruation is much worse, pain and less sleep..."* (Ella). |

6. The COMIP experience was heavily intertwined, and it varied between individuals which primary factor was central to their experience

All the participants experienced the four primary COMIP factors; however, it varied between individuals which primary factor was central to their experience. For some it was one of the four COMIP factors and for others it was a combination of more than one COMIP factor. For Tristan the most important part of COMIP was lack of energy, a daytime symptom and for Ella it was stress. To provide further insight into the intertwined nature of the primary COMIP factors Table 5 provides descriptions of how they affected each other.

**Table 5.**
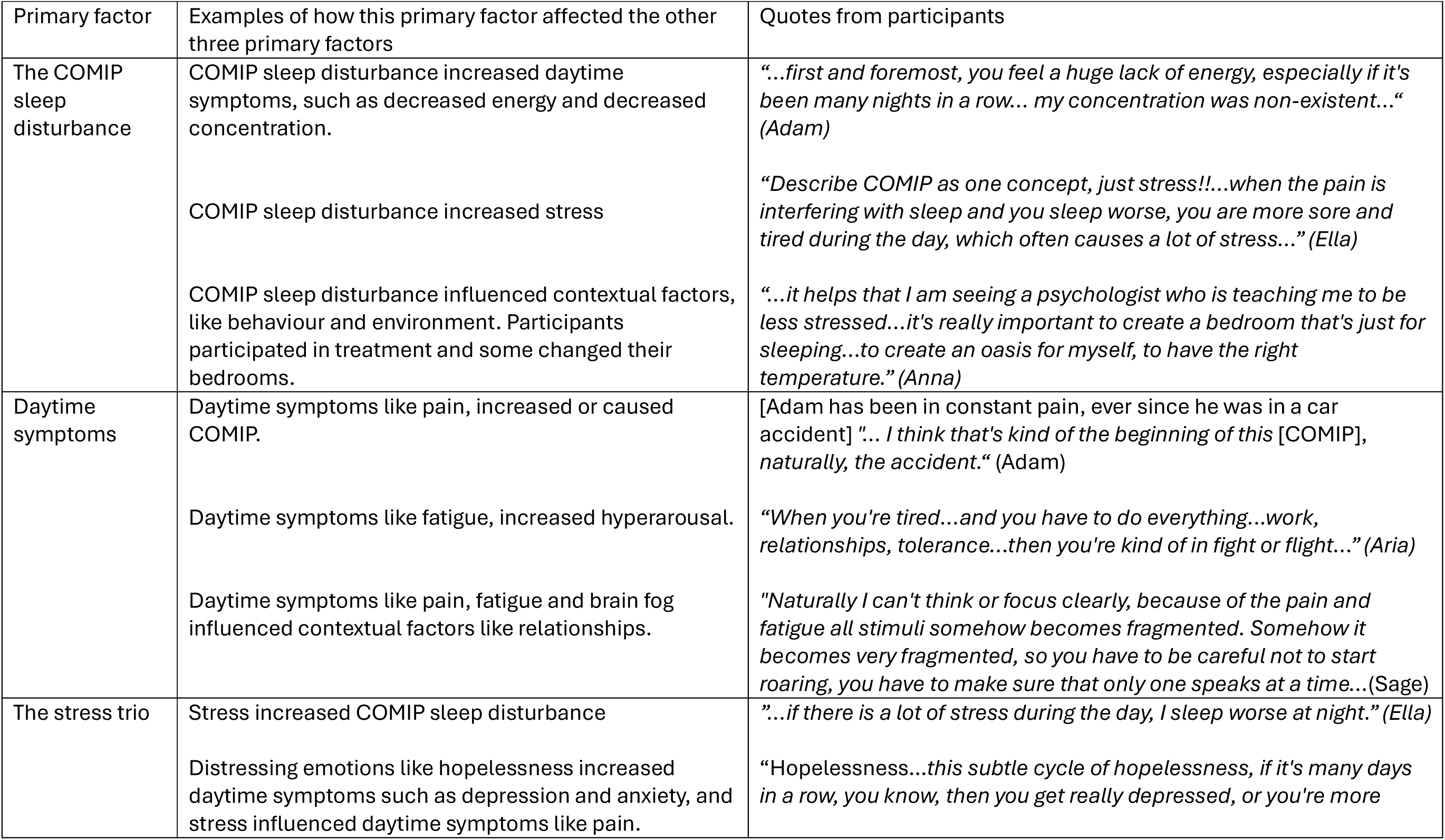
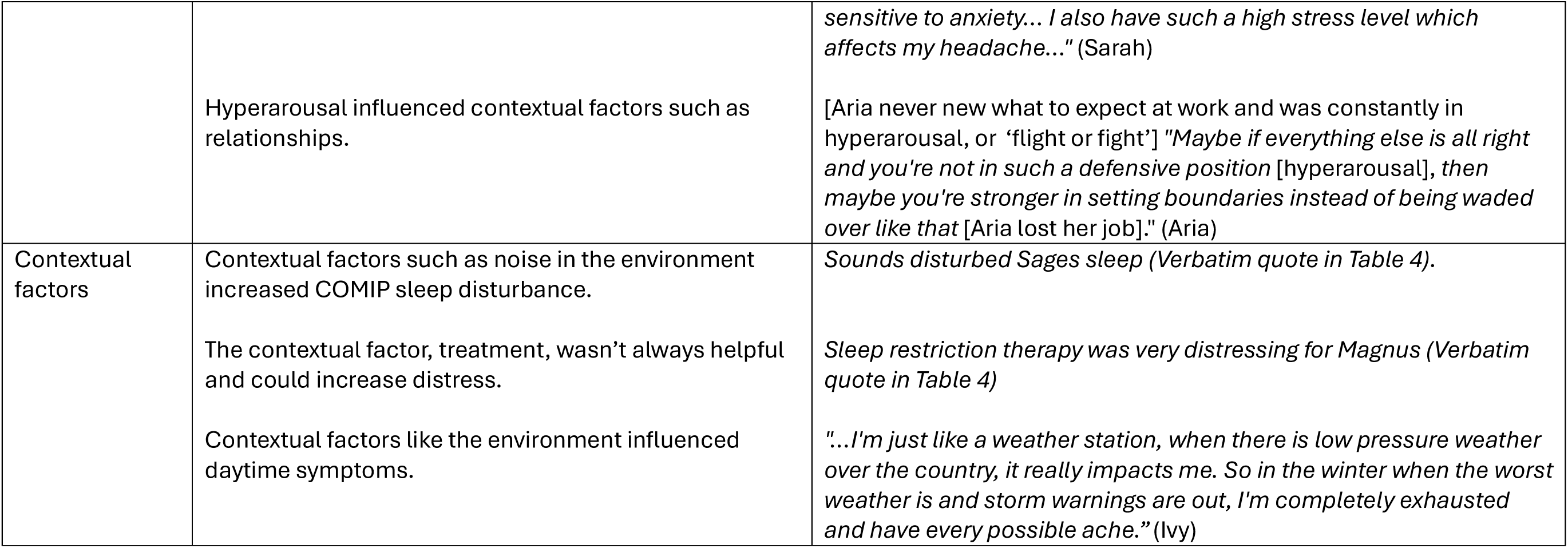
Examples of how the primary factors affected each other with quotes from participants.

| Primary factor | Examples of how this primary factor affected the other three primary factors | Quotes from participants |
| --- | --- | --- |
| The COMIP sleep disturbance | COMIP sleep disturbance increased daytime symptoms, such as decreased energy and decreased concentration.<br>COMIP sleep disturbance increased stress<br>COMIP sleep disturbance influenced contextual factors, like behaviour and environment. Participants participated in treatment and some changed their bedrooms. | *"...first and foremost, you feel a huge lack of energy, especially if it's been many nights in a row... my concentration was non-existent..." (Adam)*<br>*"Describe COMIP as one concept, just stress!!...when the pain is interfering with sleep and you sleep worse, you are more sore and tired during the day, which often causes a lot of stress..." (Ella)*<br>*"...it helps that I am seeing a psychologist who is teaching me to be less stressed...it's really important to create a bedroom that's just for sleeping...to create an oasis for myself, to have the right temperature." (Anna)* |
| Daytime symptoms | Daytime symptoms like pain, increased or caused COMIP.<br>Daytime symptoms like fatigue, increased hyperarousal.<br>Daytime symptoms like pain, fatigue and brain fog influenced contextual factors like relationships. | *[Adam has been in constant pain, ever since he was in a car accident] "... I think that's kind of the beginning of this [COMIP], naturally, the accident." (Adam)*<br>*"When you're tired...and you have to do everything...work, relationships, tolerance...then you're kind of in fight or flight..." (Aria)*<br>*"Naturally I can't think or focus clearly, because of the pain and fatigue all stimuli somehow becomes fragmented. Somehow it becomes very fragmented, so you have to be careful not to start roaring, you have to make sure that only one speaks at a time...(Sage)* |
| The stress trio | Stress increased COMIP sleep disturbance<br>Distressing emotions like hopelessness increased daytime symptoms such as depression and anxiety, and stress influenced daytime symptoms like pain. | *"...if there is a lot of stress during the day, I sleep worse at night." (Ella)*<br>*"Hopelessness...this subtle cycle of hopelessness, if it's many days in a row, you know, then you get really depressed, or you're more* |
|  | Hyperarousal influenced contextual factors such as relationships. | *sensitive to anxiety... I also have such a high stress level which affects my headache..." (Sarah)*<br>*[Aria never new what to expect at work and was constantly in hyperarousal, or 'flight or fight'] "Maybe if everything else is all right and you're not in such a defensive position [hyperarousal], then maybe you're stronger in setting boundaries instead of being waded over like that [Aria lost her job]."* (Aria) |
| Contextual factors | Contextual factors such as noise in the environment increased COMIP sleep disturbance.<br>The contextual factor, treatment, wasn't always helpful and could increase distress.<br>Contextual factors like the environment influenced daytime symptoms. | *Sounds disturbed Sages sleep (Verbatim quote in Table 4).*<br>*Sleep restriction therapy was very distressing for Magnus (Verbatim quote in Table 4)*<br>*"...I'm just like a weather station, when there is low pressure weather over the country, it really impacts me. So in the winter when the worst weather is and storm warnings are out, I'm completely exhausted and have every possible ache."* (Ivy) |

## Discussion

To our knowledge this is the first study with the primary aim of exploring adults lived experiences of the core and the meaning of COMIP, alongside conceptualizing the phenomenon. Before commencing the study, we had the pre-understanding that the COMIP experience is characterized by a vicious cycle of insomnia, pain and hyperarousal. However, the COMIP experience was more multifaceted, and this study introduces unique mapping of the lived experiences of COMIP. The main findings of the study were that regardless of the aetiology of participants’ pain, they all described COMIP as a vicious cycle of insomnia and pain, driven by four primary factors, where each factor influenced the other factors: (I) The COMIP sleep disturbance with reduced sleep function and reduced sleep quality; (II) Daytime symptoms with reduced daytime function and reduced quality of life; (III) The stress trio with 24-hour hyperarousal, stress and distress; and (IV) Contextual factors such as treatment and environmental factors. Exactly which COMIP factor was central to the COMIP experience varied between individuals; for some, it was one of the four main factors, while for others it was two or more of the factors.

Rearticulation is important in the developing field of sleep medicine, where AI-driven solutions are reshaping diagnostics and management [39]. We stress the foundational importance of including the voices of people with lived experiences of sleep concepts, when developing and defining those concepts. The essence of sleep itself was studied in a Dutch phenomenological study, which included 13 adults without sleep disorders [40]. Comfort and safety were identified in the study as essential for sleep and the experience of sleep was described as: *“an intertwined experience affected by and affecting other occupations”* [40] p.617, which aligns with our results. However, participants in the Dutch study experienced sleep as a recharging process for energy and recovery while our participants experienced the opposite, their sleep had turned into its inverse, it was accompanied by suffering and not rest. Our participants described their body attacking “this function” referring to the ability to sleep. Interestingly, a qualitative interview study on pain [41] found participants descriptions of incapacitating pain experiences to be similar to accounts of torture and stated that pain experiences have the potential to overwhelm basic function. In our study the participants described how insomnia and pain disturbed sleep and caused reduced sleep function such as; difficulties falling and staying asleep and shallow non-restorative sleep, as well as reduced sleep quality. In insomnia, the sleep disturbance was mainly due to hyperarousal symptoms. Pain disturbing sleep was unanimously described as difficulties with finding comfortable sleeping positions and/or awakenings due to pain.

Participants described that the insomnia and pain could not be clearly separated, that they were entangled and magnified one another. Similarly, the qualitative studies of Harman and colleagues and Skúladóttir and colleagues, both described that their participants experienced difficulties falling and staying asleep because of pain and hyperarousal symptoms [20,21].

Initially, we sought to distinguish between the core and the meaning of COMIP. However, participants quickly indicated that these aspects were inseparable. COMIP was not solely a nighttime phenomenon but also characterized by significant daytime symptoms, including increased pain, poorer mental health, and cognitive impairments such as reduced concentration and brain fog. Participants emphasized that COMIP caused a lack of energy with effects on daytime function like work and social life, and reduced quality of life. This aligns with insomnia being a 24-hour problem [24] and musculoskeletal pain being experienced as an adversarial struggle on multiple levels [22,23]. This also aligns with the fact that daytime symptoms are part of the diagnostic criteria for insomnia [7], and reduced daytime function is part of the diagnostic criteria for chronic pain [9].

The stress trio with 24-hour hyperarousal, stress and distress, ignited or magnified the COMIP sleep disturbance, daytime symptoms and contextual factors which in turn magnified hyperarousal, stress and distress. Hyperarousal was a part of the insomnia experience and was also described as being in a constant state of “fight or flight” with everything simmering. Stress was both described as being the primary driver of COMIP and as a biproduct of the sleep problem and lack of energy. Also, distressing emotions such as suffering and hopelessness were described. It was expected that the participants were experiencing hyperarousal, and while stress and distress were not part of our pre-understanding, it was not surprising when stress and distress emerged as part of one of the main themes of COMIP. Especially since significant emotional distress is part of the diagnostic criteria in chronic pain [9]. Those results also align with a meta-analytical study where a significant association was found between insomnia symptoms and emotion dysregulation, primarily in individuals with comorbid health condition [42].

The broad theoretical framework of ICF facilitated the identification of contextual factors as one of the main themes, with the sub-themes: treatment, environment, other sleep problems, personal and social factors. This was the only main theme that could not only increase COMIP but also decrease it. A key dichotomy in contextual factors was the conflict between treatment burden vs. results. All participants had tried different treatments for their sleep problems, with varying results. Digital cognitive-behavioural therapy for insomnia is an example of this, it was helpful for one participant, while the same course (the sleep restriction part) increased the sleep problem for another participant and caused severe distress. Those results align with the results of a feasibility study for hybrid cognitive behavioural therapy for pain-related insomnia [43]. Tang’s study had a large drop-out and increased pain in participants was identified as a possible side effect of the sleep restriction component. Interestingly, a retrospective study of daytime sleepiness in insomnia with over 1000 participants, found a statistically significant increase in sleepiness after cognitive behavioural therapy for insomnia, despite significant improvements in daytime symptoms and fatigue [44]. The authors pointed out that increased sleep pressure is part of the CBT-I treatment, and that the sleepiness effect of CBT-I has in some studies been shown to be temporary and reversible [44,45]. However, although CBT-I has shown positive effects for both insomnia and pain in adults with COMIP [46,47], we concur with the suggestions of Tang and colleagues [43], that for patient safety further investigation into the possible side effects of sleep restriction therapy is needed for patients with COMIP. Therefore, we recommend that randomized controlled trials are performed assessing the treatment effects of CBT-I with and without sleep restriction for COMIP patients. Other treatments should be tested in the same manner. For some individuals with COMIP an interdisciplinary rehabilitation program with education and training, delivered both individually and in group therapy, might be the most useful intervention [21], but this needs to be tested in clinical trials as well. Samea and colleagues [42] suggested that the integration of emotion regulation skills into insomnia management could improve therapeutic outcomes. This suggestion might hold some potential; however, we offer a different perspective. Pain and insomnia have both been used as torture tools and suffering was part of the distressing emotions that our participants experienced.

Hyperarousal can be a normal biological reaction to threat, and the threat of pain and insomnia should not be underestimated. We suggest that by increasing the understanding that hyperarousal, stress and distress are indeed part of COMIP, showing empathy and understanding (emotional support and validation) and by simultaneously decreasing the threat by effectively treating COMIP, the accompanying hyperarousal could decrease. This perspective aligns with the findings of a phenomenological study on patients’ needs in rehabilitation [48] where it was observed that accidents or illness usually leads to trying to cope with existential changes, while adapting to new characteristics of life and self. The authors concluded that patients in those circumstances need emotional support and recognition of their own hierarchy of needs. Our findings additionally highlight the need to recognize a patient’s individual hierarchy of needs in COMIP, because it varied between participants which COMIP factor/s were central to their experience.

The strengths of this study include that all participants reported experiencing the four primary COMIP factors, including significant emotional distress and reduced functional ability, which are reflected in the International Association for the Study of Pain (IASP) classification of chronic pain [9]. In addition, all participants described daytime symptoms, which are included in the diagnostic criteria for insomnia [7]. This alignment strengthens the validity of the findings, while the results also contribute to existing diagnostic frameworks by providing insight from the patients’ perspective.

The study also has several limitations. The sample may not be representative of the broader COMIP population, as all participants were Icelandic, most had attended rehabilitation, and the gender distribution was skewed towards females. Further research is needed to explore the experiences of COMIP across diverse subgroups. Future research on COMIP should address several key areas. This includes investigating the origins and consequences of COMIP, as well as exploring the lived experiences of individuals with COMIP across different cultural contexts.

Further research is also needed to examine the potential adverse effects of sleep restriction in this population, evaluate differential treatment effects, and better understand the bidirectional relationship between insomnia and pain.

### Conclusion

This research offers three-fold contribution, firstly empirical accounts of living with COMIP, secondly the introduction of novel concepts and thirdly, the conceptualization of COMIP. Our findings indicate that COMIP is a 24-hour multifaceted experience. Moreover, when evaluating patients with COMIP, there is value in exploring the four COMIP factors: (i) The COMIP sleep disturbance with reduced sleep function and reduced sleep quality; (ii) Daytime symptoms with reduced daytime function and reduced quality of life; (iii) The stress trio with 24-hour hyperarousal, stress and distress; and (iv) Contextual factors like treatment and the environment. Although it varies between individuals which factor/s is central to their COMIP experience we emphasize that COMIP is distressing, and patients are likely to need emotional support and validation.

What this study adds

A new perspective, exploring COMIP as a phenomenon.

Rich descriptions of the lived experiences of COMIP.

Conceptual advancement of COMIP with the four following novel concepts emerging:

The COMIP sleep disturbance. Sleep function. The stress trio, and finally the COMIP primary factors.

## Acknowledgements

First and foremost, sincere thanks to the participants for their valuable contribution. Bryndís Elísa Árnadóttir and Hafdís Gunnarsdóttir, registered nurses at Reykjalundur rehabilitation, thank you so much for recruiting the first three participants. Finally, sincere thanks to Dr. Jónína Sigurgeirsdóttir, expert in rehabilitation for review and advice on the manuscript.

## Data availability statement

The data that supports the findings of this study are presented as part of the material in this article. The complete data supporting the findings of this study consist of verbatim transcriptions of interviews which will not be made publicly available.

## Ethics and consent

This study was approved by the National Bioethics Committee and Data Protection Agency of Iceland, approval number VSN2501010 and the chief physician at Reykjalundur rehabilitation. Participants signed an informed consent.

## Contributions

The following authors contributed to: Study design (AA, ASI, HS, SH, KAÓ, GNP and ESA). Interviews conducted and written up verbatim (AA). Triangulation and data analysis (AA, ASI, HS, EB, AV and ESA). Drafting of the manuscript (AA, ASI, HS, SH and ESA). Revision of the manuscript: All authors.

## Appendix A: Interview frame

Before commencing the interview, the researcher introduced herself and thanked the participants for their willingness to participate in the study. Then she presented the aim of the study and logistics, such as the interview being recorded and transcribed verbatim. Although there was an interview guide with guiding questions, the participants were encouraged to talk freely.

1. Approximately when did your insomnia start?
2. Can you describe your insomnia to me?
3. What is your experience with insomnia?
4. How does insomnia affect your health?
5. How does insomnia affect your life?
6. Approximately when did pain start to disturb your sleep?
7. Can you describe the pain as a sleep disturbance to me?
8. What is your experience with pain that disturbs sleep?
9. How does pain as a sleep disturbance affect your health?
10. How does pain-sleep-disturbance affect your life?
11. What is your experience with the impact of insomnia on the pain?
12. What is your experience with the impact of pain on insomnia?
13. Can you describe to me the difference between insomnia and pain-sleep-disturbance?
14. Can you describe to me the two-way relationship between insomnia and pain-sleep disturbance?
15. What is your experience with insomnia and pain-sleep-disturbance together?
16. Can you describe your insomnia and pain-sleep-disturbance to me as a concept?
17. How do insomnia and pain-sleep-disturbance together affect your health?
18. How do insomnia and pain-sleep-disturbance, together affect your life?
19. Can you tell me about the treatments you have tried for your sleep problems?
20. Which treatments helped the most with your sleep problems?
21. Which treatments helped the least with your sleep problems?
22. Is there anything else you would like to tell me?

